# Comparative efficacy of pharmacological and non-pharmacological adjunctive treatments for prominent or persistent negative symptoms of schizophrenia: a systematic review and network meta-analysis

**DOI:** 10.64898/2026.07.29.26359223

**Authors:** Chenkai Yangyang, Jia Chen, Xiaoqiang Xiao, Yifan Li, Hongjun Du, Wenjiao Min, Xu Zhang

## Abstract

**Background:** Negative symptoms are persistent determinants of disability in schizophrenia and often respond incompletely to antipsychotic treatment.

**Objective:** To compare the efficacy of pharmacological and non-pharmacological add-on treatments for prominent or persistent negative symptoms using network meta-analysis.

**Methods:** This systematic review followed PRISMA 2020 and PRISMA-NMA and was registered in PROSPERO (CRD420261422218). PubMed, Europe PMC, Semantic Scholar and Crossref were searched from inception to 29 July 2026, supplemented by citation chasing. Eligible studies were randomized controlled trials in adults with DSM/ICD schizophrenia or schizoaffective disorder, prominent or persistent negative symptoms, stable antipsychotic treatment and an adjunctive intervention. Outcomes were Positive and Negative Syndrome Scale negative subscale or Scale for the Assessment of Negative Symptoms scores. Random-effects frequentist networks estimated standardized mean differences (SMDs) with 95% confidence intervals (CIs); negative values favoured add-on treatment. Risk of bias was assessed with Cochrane RoB 2.

**Results:** Forty-nine unique RCTs met the clinical and design criteria, of which 28 (2,067 randomized; 1,933 analysable participants) contributed to the locked quantitative dataset. The primary connected network included 24 trials, 25 treatments and 31 comparison estimates. Fourteen add-ons had CIs excluding the null versus a broad control node. The highest P-scores were observed for mirtazapine (SMD −2.38, 95% CI −3.55 to −1.21), granisetron (−1.96, −2.74 to −1.17), tropisetron (−1.82, −2.59 to −1.05), minocycline (−1.78, −2.55 to −1.01) and memantine (−1.54, −2.28 to −0.80). Heterogeneity was low (τ = 0.119; τ ² = 0.014), but the predominantly star-shaped network had zero inconsistency degrees of freedom. Overall RoB 2 judgements were low for eight trials, some concerns for 15 and high for five.

**Conclusions:** Several pharmacological and non-pharmacological add-ons showed potentially important efficacy signals. Because most nodes were informed by single small trials, direct active comparisons were scarce, inconsistency could not be evaluated and risk-of-bias concerns were common, the treatment hierarchy should be considered hypothesis-generating rather than a basis for firm clinical recommendations.

Registration: PROSPERO CRD420261422218

**Key Points:**

- This review compares pharmacological and non-pharmacological adjuncts in adults selected for prominent or persistent negative symptoms while receiving stable antipsychotic medication.
- Mirtazapine, granisetron, tropisetron, minocycline and memantine had the highest P-scores, while tDCS, rTMS and body-oriented psychotherapy also showed efficacy signals versus broad control.
- The evidence network was sparse and star-shaped, most interventions were supported by one small trial, and inconsistency was not estimable; rankings therefore require cautious interpretation.

## Introduction

Negative symptoms—including blunted affect, alogia, avolition, asociality and anhedonia—are core features of schizophrenia and major determinants of impaired social and occupational functioning [1–3]. Contemporary frameworks distinguish primary negative symptoms from secondary phenomena related to positive symptoms, depression, extrapyramidal adverse effects, sedation, substance use or environmental deprivation [2–6]. This distinction is clinically important because secondary symptoms may improve when their cause is treated, whereas persistent primary symptoms remain a substantial unmet therapeutic need [4–6].

Antipsychotic drugs are essential for relapse prevention and control of positive symptoms, but their direct effects on persistent negative symptoms are limited and difficult to separate from improvements in other symptom domains [5, 9]. Adjunctive strategies have therefore targeted serotonergic, glutamatergic, cholinergic, inflammatory and neuroplastic pathways, while non-pharmacological approaches have included cognitive and body-oriented therapies, exercise and non-invasive brain stimulation. Earlier meta-analyses identified signals across several intervention classes, but clinical interpretability was constrained by heterogeneous populations, outcome scales and broad inclusion criteria [7, 8].

Conventional pairwise syntheses cannot provide a coherent comparison when many adjuncts have been evaluated almost exclusively against placebo or sham control. Network meta-analysis can combine direct and indirect evidence, provided the transitivity assumption is plausible, and can describe a relative hierarchy without implying that rankings alone establish certainty or clinical importance. We therefore conducted a systematic review and frequentist network meta-analysis of randomized controlled trials (RCTs) enrolling adults with schizophrenia or schizoaffective disorder, prominent or persistent negative symptoms and stable background antipsychotic treatment. We aimed to estimate the relative efficacy of pharmacological and non-pharmacological add-on interventions and to examine network geometry, heterogeneity, risk of bias and robustness.

## Methods

### 2.1 Protocol, registration and reporting

The review was registered in PROSPERO (CRD420261422218) and is reported according to PRISMA 2020 and the PRISMA extension for network meta-analysis [10–12]. The review question, eligibility criteria, principal outcome and random-effects network approach were defined before quantitative synthesis.

The registered plan included Embase and Cochrane CENTRAL. On the search dates, neither source provided a verified, exportable session through an authorized institutional route; an unstable third-party Embase proxy was not used for data export. Europe PMC, Semantic Scholar Academic Graph, Crossref and structured citation chasing were added to reduce the risk of missed studies. This change was documented as a protocol deviation and is considered when interpreting review completeness.

An amendment to the PROSPERO record was initiated on 29 July 2026 to revise the title so that it better reflects the final scope of the analysis. The registration number (CRD420261422218) remains unchanged. This proposed administrative title update does not alter the review question, eligibility criteria, outcomes or planned analytical framework; the search-source deviation described above was documented separately.

### 2.2 Information sources and search strategy

PubMed/MEDLINE was searched from inception to 27 July 2026 and Europe PMC from inception to 28 July 2026. Semantic Scholar Academic Graph and Crossref were searched on 29 July 2026. We also examined the trial list and supplementary material of a recent comprehensive review [8] and checked references of eligible reports. Searches combined terms for schizophrenia or schizoaffective disorder, negative symptoms or relevant scales, and randomized, placebo-controlled or blinded trials. No language or publication year restriction was applied.

The PubMed strategy was: (“Schizophrenia”[Mesh] OR “Schizoaffective Disorder”[Mesh] OR schizophreni*[Title/Abstract] OR schizoaffective*[Title/Abstract]) AND (“negative symptom”[Title/Abstract] OR “negative symptoms”[Title/Abstract] OR “deficit syndrome”[Title/Abstract] OR “persistent negative”[Title/Abstract] OR “predominant negative”[Title/Abstract] OR “prominent negative”[Title/Abstract] OR “PANSS negative”[Title/Abstract] OR “PANSS-N”[Title/Abstract] OR SANS[Title/Abstract] OR “Scale for the Assessment of Negative Symptoms”[Title/Abstract]) AND (randomized controlled trial[Publication Type] OR controlled clinical trial[Publication Type] OR randomized[Title/Abstract] OR randomised[Title/Abstract] OR placebo[Title/Abstract] OR blind*[Title/Abstract] OR mask*[Title/Abstract]) NOT (animals[MeSH Terms] NOT humans[MeSH Terms]). Source-specific translations and all supplementary queries are retained in the search audit files.

Semantic Scholar returned 6,738 records; because 4,250 lacked a visible abstract and some metadata were unreliable, 2,553 records containing all three visible concepts (schizophrenia, negative symptoms and trial design) were retained before deduplication. Eight Crossref relevance queries returned up to 1,000 records each; the same local three-concept filter retained 363 unique records. These sources were treated as supplementary discovery tools, not as substitutes equivalent to Embase or CENTRAL.

### 2.3 Eligibility criteria

We included parallel-group or randomized-withdrawal RCTs with at least 10 participants. Participants were adults aged 18 years or older with schizophrenia or schizoaffective disorder diagnosed using DSM or ICD criteria. Studies had to prospectively select participants for prominent, predominant, persistent or primary negative symptoms at a criterion level (for example, PANSS negative-subscale scores around 20 or an explicitly defined equivalent construct) and to use stable antipsychotic treatment before randomization.

Eligible interventions were pharmacological or non-pharmacological treatments added to ongoing antipsychotic care. Pharmacological classes included glutamatergic, serotonergic, anti-inflammatory, neuropeptide, cholinergic, nutritional and other agents. Non-pharmacological interventions included cognitive remediation, cognitive behavioural therapy, mindfulness, repetitive transcranial magnetic stimulation (rTMS), transcranial direct current stimulation (tDCS), exercise and arts/body-oriented therapies. Comparators were placebo, sham, treatment as usual, supportive care or an eligible active adjunct.

The prespecified outcome was negative-symptom severity measured with the Positive and Negative Syndrome Scale negative subscale (PANSS-N) or the Scale for the Assessment of Negative Symptoms (SANS) at the end of the randomized phase. A directly reported modified PANSS negative-symptom score was retained only in a disconnected active-control component. We excluded samples dominated by another primary mental disorder, severe medical or neurological illness or active substance misuse; non-adjunctive treatment; non-random allocation; non-independent or secondary reports without the target outcome; unusable numerical data; duplicate publications; and total sample size below 10.

### 2.4 Study selection and report linkage

Records were deduplicated hierarchically by exact PMID, normalized DOI, and normalized title plus year, followed by manual review of near-duplicate titles. Screening was conducted in two structured passes using a PICOS form. The review team rechecked all potentially eligible reports and all full-text exclusions; disagreements were resolved by consensus. Multiple reports from the same randomized trial were linked using author, sample, intervention, trial-registration and recruitment details, and one principal efficacy report was retained for citation and extraction.

### 2.5 Data extraction and outcomes

A standardized workbook captured study design, country, diagnostic criteria, negative-symptom entry criterion, antipsychotic stability, intervention and comparator, randomized and analysed sample sizes, duration, outcome scale, endpoint or change-score means, standard deviations and analysis population. One extraction was verified against the full report in a second pass, with discrepancies resolved by consensus. For trials reporting several time points, we used the end of the randomized treatment phase. Change scores were preferred when complete; otherwise endpoint scores were used and the data basis was recorded for sensitivity analysis.

The primary effect measure was the standardized mean difference (SMD) with 95% confidence interval (CI), calculated from arm-level mean, standard deviation and sample size. Negative values indicated lower (better) negative-symptom scores with the add-on treatment. Scale-specific networks used mean differences (MDs) and were not combined across PANSS-N, SANS total/composite, modified SANS or SANS mean-item metrics. Usall et al. (2016) was reserved for sensitivity analysis because change-score standard deviations were reconstructed from reported within-group 95% CIs.

### 2.6 Risk-of-bias assessment

Risk of bias for the negative-symptom outcome at the end of randomized treatment was assessed using Cochrane RoB 2 [18] for the effect of assignment to intervention. We judged bias arising from the randomization process (D1), deviations from intended interventions (D2), missing outcome data (D3), measurement of the outcome (D4), and selection of the reported result (D5). Domain judgements were low risk, some concerns or high risk; the overall judgement followed the RoB 2 algorithm. Assessments were checked against the full report and accessible trial-registration information.

### 2.7 Network meta-analysis

We fitted frequentist random-effects network meta-analyses in R version 4.6.0 using netmeta version 3.6-0 and meta version 8.5-0 [17, 19, 20]. Multi-arm trials retained a common study identifier so that within-trial correlation was handled by the network model. The primary network pooled placebo, sham stimulation, supportive counselling and treatment as usual into a broad Control node. A sensitivity analysis retained granular comparator and dose nodes.

A common between-study variance was estimated by restricted maximum likelihood. We report τ, τ² and Cochran’s Q [13]. Network connectivity was checked before every model; disconnected components were analysed separately and were not assigned a common ranking. Relative rankings used random-effects P-scores with smaller outcome values defined as desirable [14]. P-scores were interpreted descriptively and never as certainty, safety or clinical-importance measures. Statistical significance was defined by a 95% CI excluding zero.

We planned design-by-treatment decomposition and local node splitting where the network contained loops and residual inconsistency degrees of freedom [12, 15]. These procedures were not forced when the network structure made them non-identifiable. Transitivity was assessed clinically by comparing diagnosis, negative-symptom enrichment, background antipsychotic stability, outcome timing and comparator type across comparisons.

### 2.8 Sensitivity analyses and reporting-bias assessment

Sensitivity analyses added the study with reconstructed standard deviations; excluded trials flagged for completer-only data or PICOS sensitivity; separated change-score from endpoint-score studies; disaggregated comparator and dose nodes; and fitted scale-specific PANSS-N, SANS total/composite and modified-SANS networks. The cognitive behavioural therapy versus cognitive remediation comparison was reported as a disconnected direct component.

For the primary connected network, small-study and reporting-bias signals were explored using a comparison-adjusted funnel plot and Egger regression [16]. Because funnel asymmetry in a network can also reflect heterogeneity, multi-arm correlation and network geometry, this analysis was considered exploratory. A formal certainty-of-evidence framework was not applied; risk of bias, precision, heterogeneity, transitivity and network geometry were considered qualitatively when interpreting the estimates.

## Results

### 3.1 Study selection

The searches yielded 10,055 records: 3,369 from PubMed, 3,770 from Europe PMC, 2,553 Semantic Scholar records retained after local filtering and 363 Crossref records retained after local filtering. After removal of 5,352 duplicates, 4,703 records were screened and 4,464 were excluded at title or abstract stage. Of 239 reports sought for detailed assessment, 23 could not be retrieved or contained insufficient information to establish eligibility. We assessed 216 reports and excluded 167: population (n = 94), not an independent primary report (n = 33), study design (n = 20), other PICOS mismatch (n = 11), outcome (n = 5), intervention/background treatment (n = 3) and retraction (n = 1). Forty-nine unique RCTs met the clinical and design criteria [21–69]. Twenty-eight contributed to the locked quantitative dataset; the other 21 did not contribute because usable outcome data were not present in the assembled extraction set (Fig. 1).

**Fig. 1.**
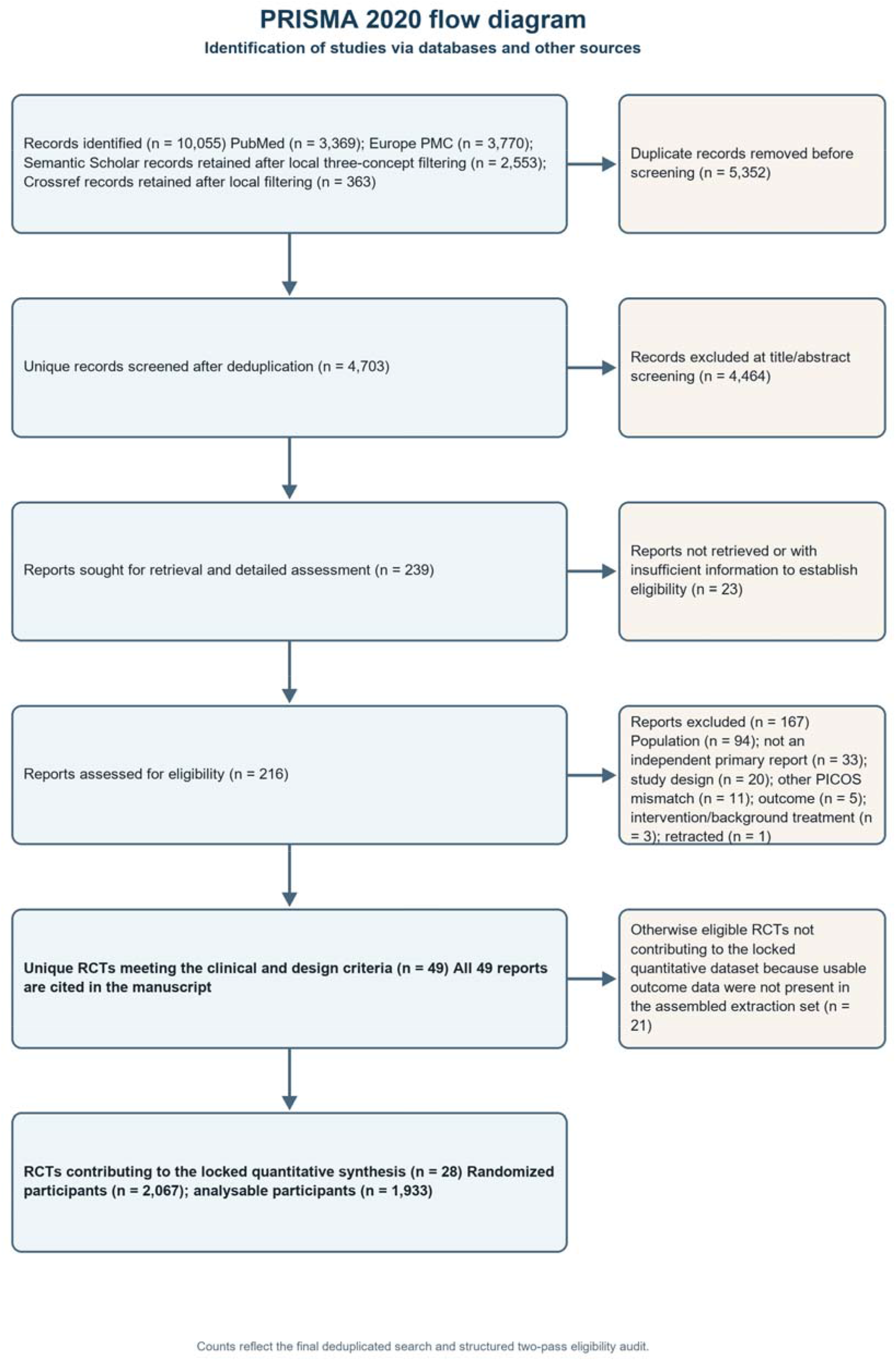
PRISMA 2020 flow diagram. Semantic Scholar and Crossref counts are records retained after the documented local three-concept filters. Twenty-one RCTs met the clinical and design criteria but did not contribute to the locked quantitative synthesis because usable outcome data were not present in the assembled extraction set

### 3.2 Study and evidence characteristics

The locked analytic dataset comprised 28 RCTs, including 2,067 randomized participants and 1,933 participants represented in the analysable data, and contributed 32 observed contrasts. Twenty-seven trials formed the prespecified main-analysis set; Usall et al. (2016) was reserved for sensitivity analysis because change-score standard deviations were reconstructed from within-group 95% CIs. Twenty-two trials evaluated pharmacological add-ons and six evaluated non-pharmacological add-ons. Three trials used multi-arm designs, and treatment duration ranged from 3 to 52 weeks (Table 1).

**Table 1.**
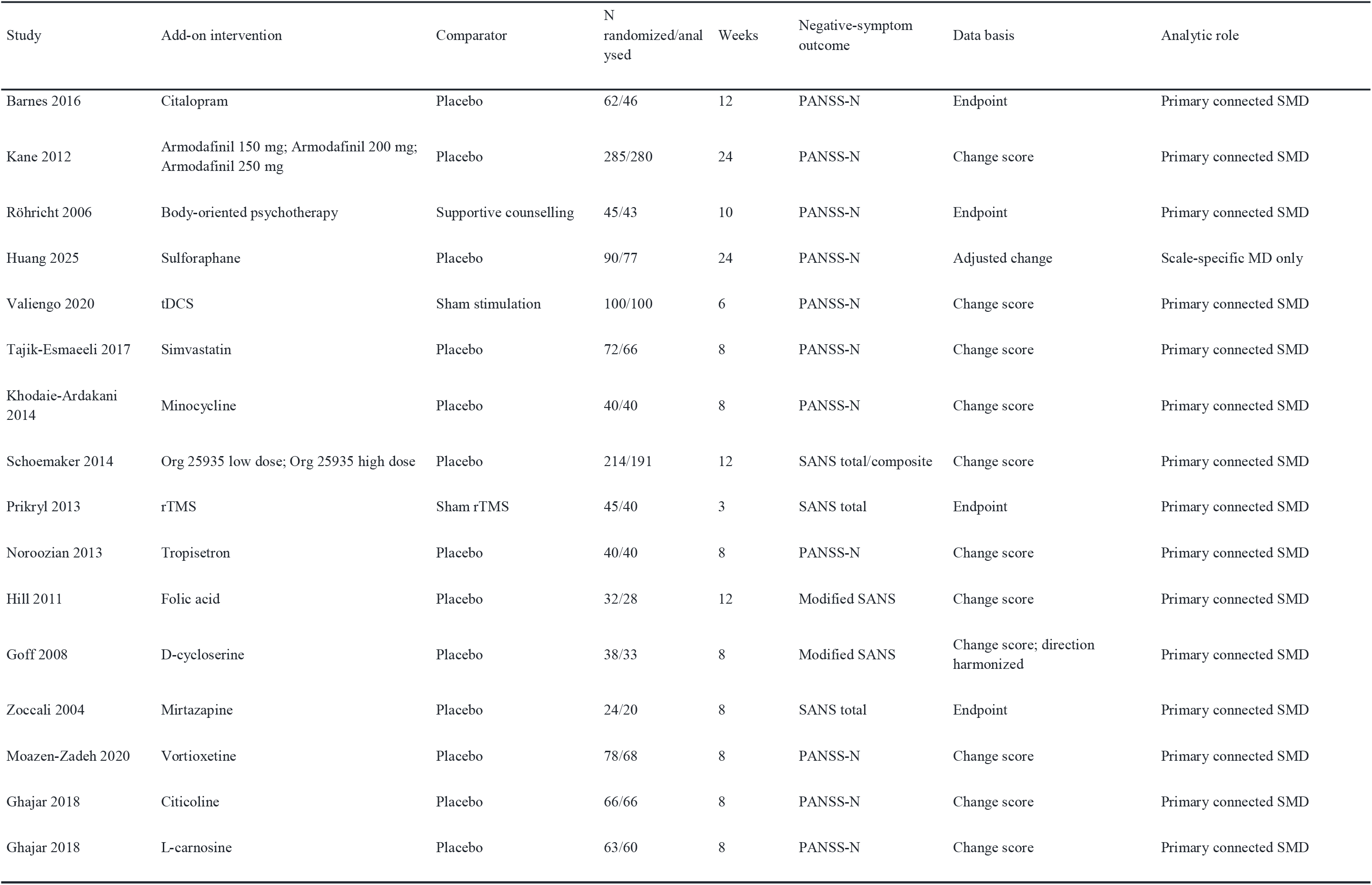
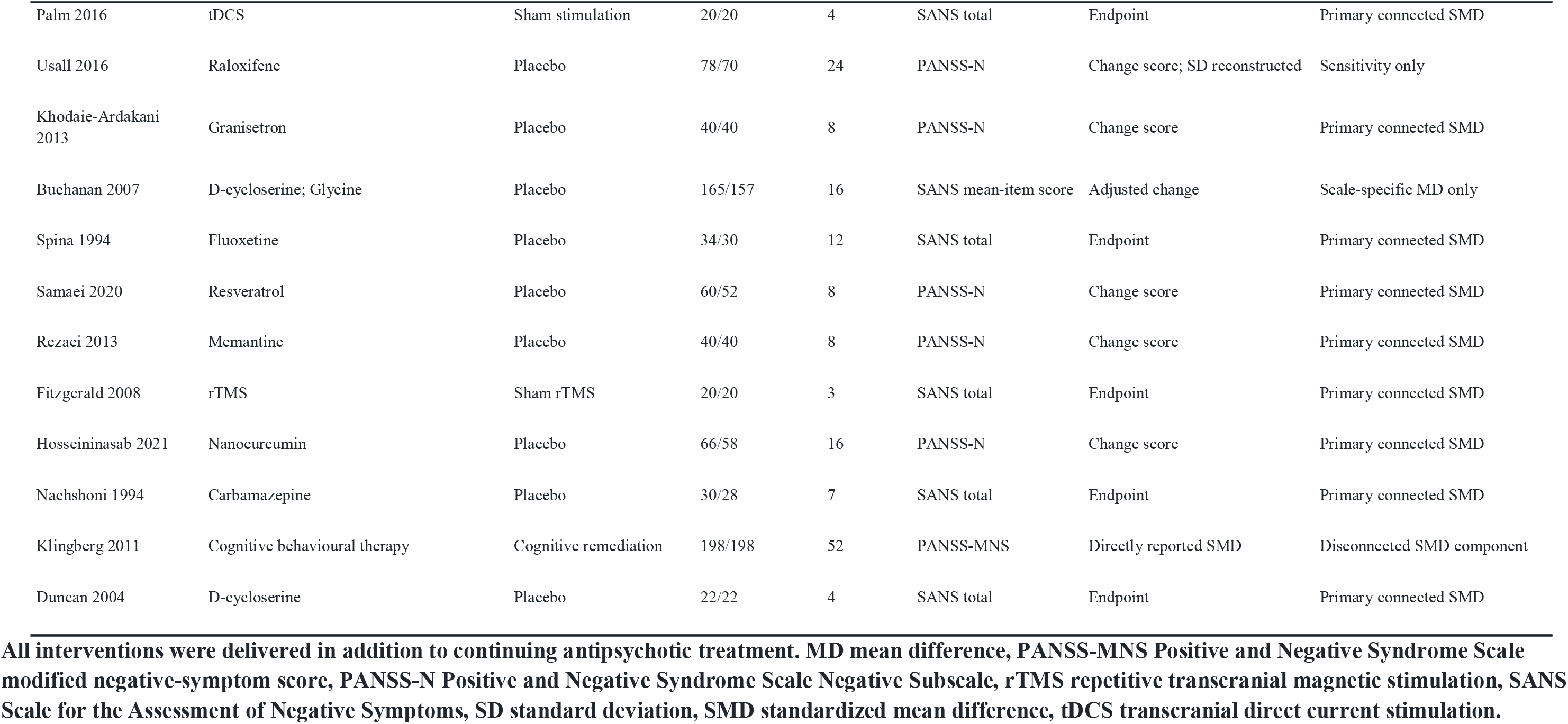
Characteristics of randomized controlled trials contributing analysable data. All interventions were delivered in addition to continuing antipsychotic treatment. MD mean difference, PANSS-MNS Positive and Negative Syndrome Scale modified negative-symptom score, PANSS-N Positive and Negative Syndrome Scale Negative Subscale, rTMS repetitive transcranial magnetic stimulation, SANS Scale for the Assessment of Negative Symptoms, SD standard deviation, SMD standardized mean difference, tDCS transcranial direct current stimulation.

PANSS-N data were available from 16 trials (15 in the main set), SANS total or composite scores from eight trials, modified-SANS scores from two trials, and a SANS mean-item score from one trial. Cross-scale SMDs were available for 25 main-set trials: 24 supplied arm-level means, standard deviations and sample sizes, whereas Klingberg et al. (2011) supplied a directly reported standardized effect. Sulforaphane (Huang et al. 2025) and the D-cycloserine/glycine trial (Buchanan et al. 2007) could not be standardized from the available data and therefore contributed only to scale-specific MD analyses.

### 3.3 Network geometry

The primary connected network contained 24 studies, 25 treatment nodes, 31 direct pairwise comparison estimates and 21 designs (Fig. 2). The geometry was predominantly star-shaped around the broad Control node, which combined placebo, sham stimulation, supportive counselling and treatment-as-usual comparators. Multi-arm armodafinil and Org 25935 trials supplied the principal active–active connections. Cognitive behavioural therapy and cognitive remediation formed a separate two-node component; their direct comparison did not show a clear difference (SMD 0.12, 95% CI −0.16 to 0.40) and was excluded from the primary treatment hierarchy.

**Fig. 2.**
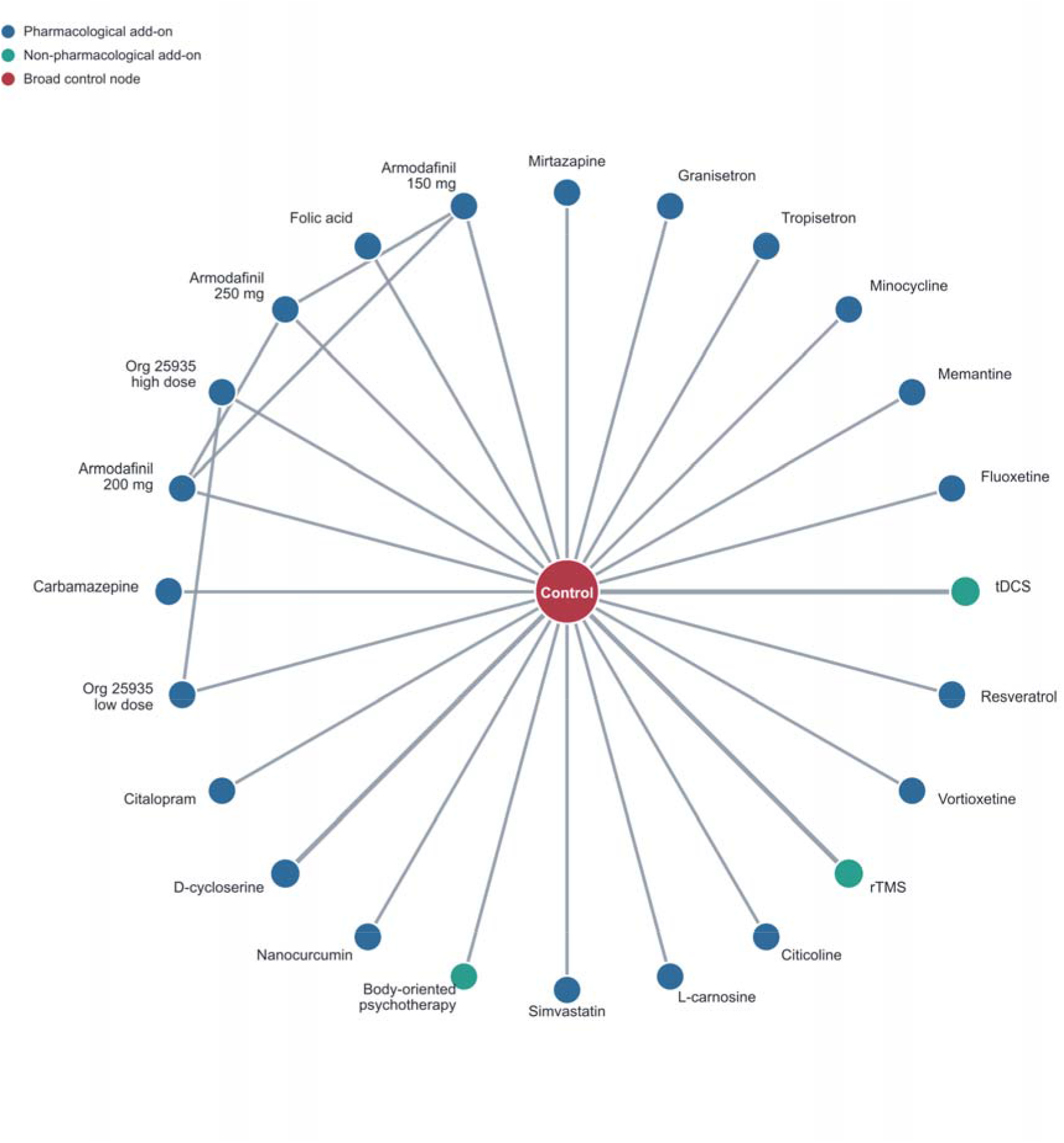
Network geometry of the primary connected SMD analysis. The network contained 24 studies, 25 treatment nodes and 31 direct pairwise comparison estimates. Node size reflects the number of contributing studies and line width reflects the number of direct comparisons. The disconnected cognitive behavioural therapy–cognitive remediation component is not shown

### 3.4 Primary network meta-analysis

Fourteen add-on interventions had 95% CIs that excluded the null in comparison with Control (Table 2; Fig. 3). The five highest-ranked treatments were mirtazapine (SMD −2.38, 95% CI −3.55 to −1.21; P-score 0.958), granisetron (−1.96, −2.74 to −1.17; 0.916), tropisetron (−1.82, −2.59 to −1.05; 0.891), minocycline (−1.78, −2.55 to −1.01; 0.883) and memantine (−1.54, −2.28 to −0.80; 0.827). Fluoxetine, tDCS, resveratrol, vortioxetine, rTMS, citicoline, L-carnosine, simvastatin and body-oriented psychotherapy also favoured active treatment, with SMDs ranging from −1.31 to −0.75.

**Fig. 3.**
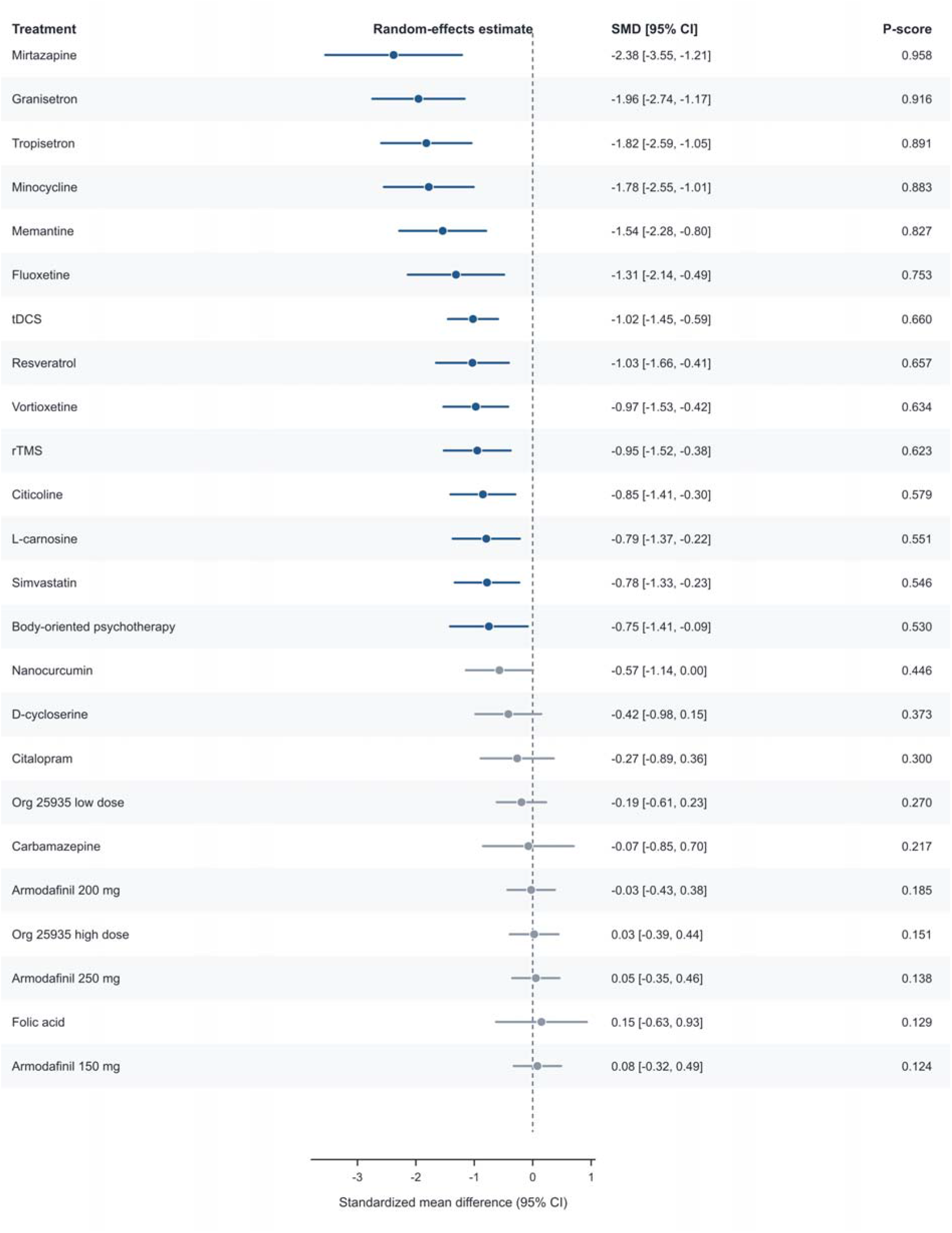
Primary random-effects network estimates versus the broad Control node. Negative SMD values favour the add-on intervention. Navy symbols denote 95% CIs excluding the null; grey symbols denote intervals crossing the null. P-scores are descriptive and should be interpreted with the precision and evidence base for each treatment

**Table 2.** Primary random-effects network estimates versus Control. Negative SMD values favour the add-on treatment. P-scores are descriptive rankings and do not incorporate certainty of evidence, safety or clinical importance. CI confidence interval, SMD standardized mean difference.

| Treatment | SMD | Lower 95% CI | Upper 95% CI | p value | P-score | Rank |
| --- | --- | --- | --- | --- | --- | --- |
| Mirtazapine | -2.38 | -3.55 | -1.21 | <0.001 | 0.958 | 1 |
| Granisetron | -1.96 | -2.74 | -1.17 | <0.001 | 0.916 | 2 |
| Tropisetron | -1.82 | -2.59 | -1.05 | <0.001 | 0.891 | 3 |
| Minocycline | -1.78 | -2.55 | -1.01 | <0.001 | 0.883 | 4 |
| Memantine | -1.54 | -2.28 | -0.80 | <0.001 | 0.827 | 5 |
| Fluoxetine | -1.31 | -2.14 | -0.49 | 0.002 | 0.753 | 6 |
| tDCS | -1.02 | -1.45 | -0.59 | <0.001 | 0.660 | 7 |
| Resveratrol | -1.03 | -1.66 | -0.41 | 0.001 | 0.657 | 8 |
| Vortioxetine | -0.97 | -1.53 | -0.42 | <0.001 | 0.634 | 9 |
| rTMS | -0.95 | -1.52 | -0.38 | 0.001 | 0.623 | 10 |
| Citicoline | -0.85 | -1.41 | -0.30 | 0.003 | 0.579 | 11 |
| L-carnosine | -0.79 | -1.37 | -0.22 | 0.007 | 0.551 | 12 |
| Simvastatin | -0.78 | -1.33 | -0.23 | 0.005 | 0.546 | 13 |
| Body-oriented psychotherapy | -0.75 | -1.41 | -0.09 | 0.027 | 0.530 | 14 |
| Nanocurcumin | -0.57 | -1.14 | 0.00 | 0.052 | 0.446 | 15 |
| D-cycloserine | -0.42 | -0.98 | 0.15 | 0.147 | 0.373 | 16 |
| Citalopram | -0.27 | -0.89 | 0.36 | 0.407 | 0.300 | 17 |
| Org 25935 low dose | -0.19 | -0.61 | 0.23 | 0.373 | 0.270 | 18 |
| Carbamazepine | -0.07 | -0.85 | 0.70 | 0.851 | 0.217 | 19 |
| Armodafinil 200 mg | -0.03 | -0.43 | 0.38 | 0.896 | 0.185 | 20 |
| Org 25935 high dose | 0.03 | -0.39 | 0.44 | 0.906 | 0.151 | 21 |
| Armodafinil 250 mg | 0.05 | -0.35 | 0.46 | 0.794 | 0.138 | 22 |
| Folic acid | 0.15 | -0.63 | 0.93 | 0.708 | 0.129 | 23 |
| Armodafinil 150 mg | 0.08 | -0.32 | 0.49 | 0.696 | 0.124 | 24 |

**Table 3.** Primary network diagnostics and small-study-effect assessment. Global and local inconsistency could not be estimated because the network had zero inconsistency degrees of freedom. The comparison-adjusted funnel analysis is exploratory. REML restricted maximum likelihood.

| Domain | Metric | Value | Comment |
| --- | --- | --- | --- |
| Network size | Studies | 24 |  |
| Network size | Treatments | 25 |  |
| Network size | Pairwise comparison estimates | 31 |  |
| Network size | Designs | 21 |  |
| Heterogeneity | $\tau$ | 0.119 | REML |
| Heterogeneity | $\tau^2$ | 0.014 | REML |
| Cochran Q | Q (df) | 3.373 (3) | $p = 0.338$ |
| Inconsistency | Degrees of freedom | 0 | Not estimable |
| Funnel asymmetry | Egger t (df) | -0.96 (25) | $p = 0.346$ |

The estimate for nanocurcumin was close to, but did not exclude, the null (SMD −0.57, 95% CI −1.14 to 0.00; p = 0.052). D-cycloserine, citalopram, both Org 25935 dose nodes, carbamazepine, all three armodafinil doses and folic acid did not show statistically detectable differences from Control. The complete, direction-corrected active–active league table is provided in Online Resource 1.

### 3.5 Heterogeneity and inconsistency

Between-study heterogeneity in the primary model was low (τ = 0.119; τ² = 0.014). Cochran’s Q was 3.373 (3 df; p = 0.338). The network had zero inconsistency degrees of freedom; consequently, neither a global inconsistency decomposition nor local node-splitting estimates were identifiable. This non-estimability should not be interpreted as evidence that transitivity or consistency was satisfied.

### 3.6 Sensitivity and scale-specific analyses

Adding Usall et al. (2016) expanded the connected network to 25 studies and 26 treatment nodes without changing the estimates or ordering of the six highest-ranked primary treatments; raloxifene favoured active treatment by a small margin in this sensitivity model (SMD −0.54, 95% CI −1.07 to −0.01). Excluding flagged studies reduced the network to 20 studies and 21 nodes, while mirtazapine, granisetron, tropisetron, minocycline and memantine remained the five highest-ranked treatments. Because removed or added nodes were predominantly leaves of the broad-control network, estimates for remaining nodes were unchanged.

Analyses stratified by outcome-data basis supported the direction of the primary findings but were substantially sparser. In the 14-study change-score network, granisetron was highest ranked (SMD −1.96, 95% CI −2.71 to −1.20); in the 10-study endpoint-score network, mirtazapine was highest ranked (−2.38, −3.70 to −1.06). Using detailed comparator and dose nodes split the evidence into five disconnected components, precluding a single sensitivity ranking across treatments.

Scale-specific analyses gave a PANSS-N MD of −4.65 (95% CI −5.55 to −3.75) for minocycline in a 15-study network and a SANS total/composite MD of −25.80 (−42.66 to −8.94) for tDCS in an eight-study network. The two-study modified-SANS network yielded an estimate of −4.17 (−8.33 to −0.01) for D-cycloserine. Several scale-specific networks had no residual degrees of freedom and therefore could not estimate between-study heterogeneity (Table 4).

**Table 4.** Sensitivity, scale-specific and disconnected-component analyses. Leading estimates are shown only as compact component-specific summaries. They should not be compared across different scales or disconnected networks. CBT cognitive behavioural therapy, MD mean difference, PANSS-N Positive and Negative Syndrome Scale Negative Subscale, SANS Scale for the Assessment of Negative Symptoms, SMD standardized mean difference, tDCS transcranial direct current stimulation.

| Analysis | Studies | Treatments | Leading or direct estimate | Network note |
| --- | --- | --- | --- | --- |
| SMD + RCT-028 | 25 | 26 | Raloxifene: -0.54 [-1.07, -0.01] | Top six unchanged; $\tau^2 = 0.014$ |
| SMD excluding flagged studies | 20 | 21 | Mirtazapine: -2.38 [-3.55, -1.21] | Top five unchanged; $\tau^2 = 0.014$ |
| Change-score SMD | 14 | 18 | Granisetron: -1.96 [-2.71, -1.20] | Residual df = 0 |
| Endpoint-score SMD | 10 | 10 | Mirtazapine: -2.38 [-3.70, -1.06] | $\tau^2 = 0.112$; Q p = 0.205 |
| Detailed-node SMD | 25 | 32 | No global estimate | Five disconnected components |
| PANSS-N MD | 15 | 18 | Minocycline: -4.65 [-5.55, -3.75] | Residual df = 0 |
| SANS total/composite MD | 8 | 9 | tDCS: -25.80 [-42.66, -8.94] | $\tau^2 \approx 0$; Q p = 0.938 |
| Modified SANS MD | 2 | 3 | D-cycloserine: -4.17 [-8.33, -0.01] | Residual df = 0 |
| Disconnected direct SMD | 1 | 2 | CBT vs cognitive remediation: 0.12 [-0.16, 0.40] | Two-treatment component |

### 3.7 Risk of bias

For the negative-symptom outcome at the end of randomized treatment, eight of 28 trials (28.6%) were judged at low overall risk of bias, 15 (53.6%) had some concerns and five (17.9%) were at high risk (Fig. 4). Domain-level low-risk judgements were assigned to 14 trials (50.0%) for the randomization process, 26 (92.9%) for deviations from intended interventions, 12 (42.9%) for missing outcome data, 25 (89.3%) for outcome measurement and 16 (57.1%) for selection of the reported result.

**Fig. 4.**
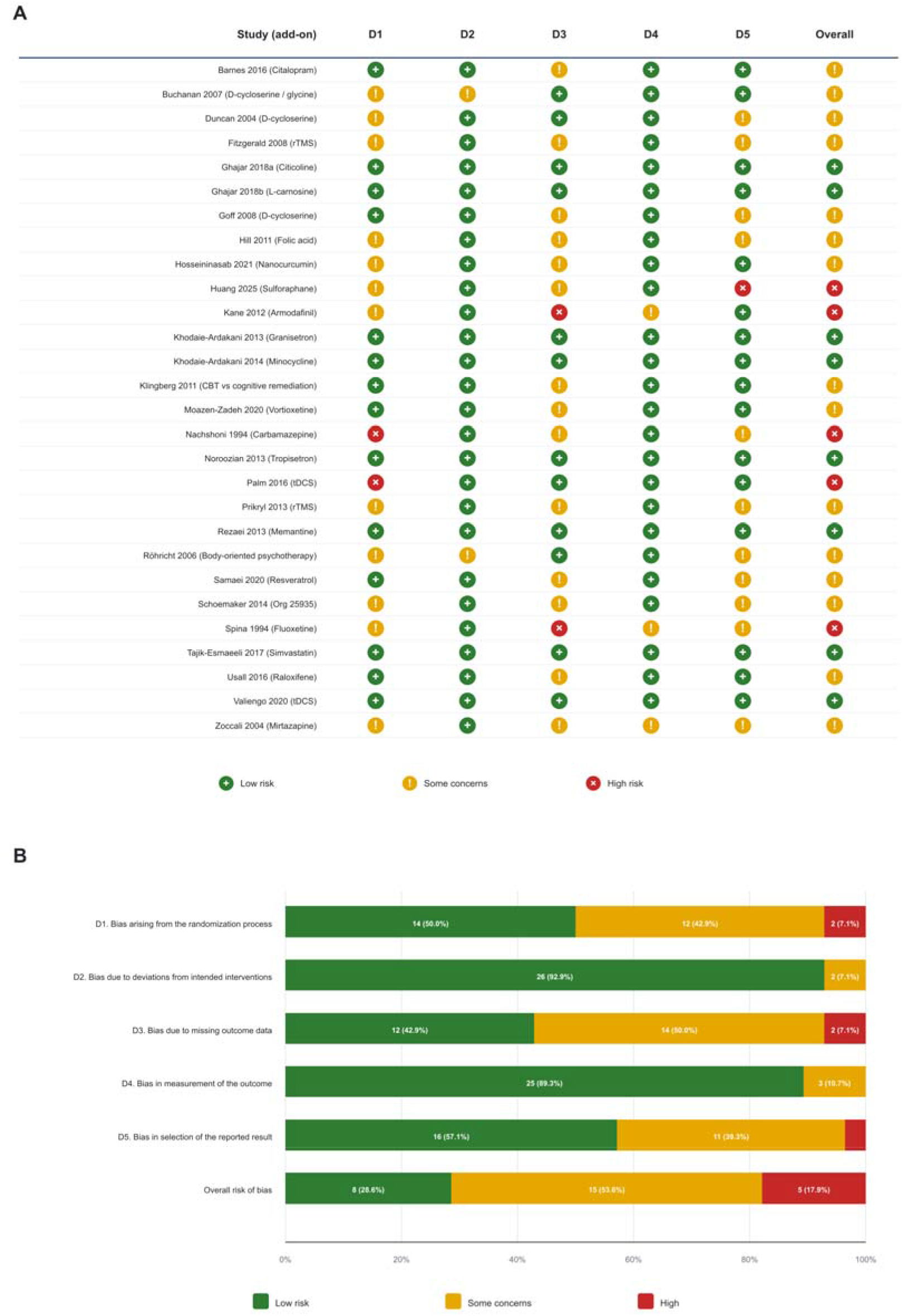
Risk of bias for the negative-symptom outcome at the end of randomized treatment. (A) Study-level Cochrane RoB 2 judgements. (B) Unweighted distribution of judgements across the five domains and overall. D1 bias arising from the randomization process, D2 bias due to deviations from intended interventions, D3 bias due to missing outcome data, D4 bias in measurement of the outcome, D5 bias in selection of the reported result

The most frequent concerns were incomplete reporting of allocation concealment, endpoint missingness handled using completer or modified intention-to-treat analyses, and absence of accessible prospective outcome or analysis specifications. High overall risk was driven by substantial, potentially outcome-related attrition in Kane et al. (2012) and Spina et al. (1994), inadequate protection of the randomized comparison in Palm et al. (2016) and Nachshoni et al. (1994), and post-completion promotion of the negative-symptom outcome in the trial registry for Huang et al. (2025).

The evidence base was not uniformly at high risk, and outcome measurement and deviations from intended interventions were generally well controlled. Nevertheless, only eight trials were low risk overall; estimates supported by single small trials or a high-risk study should be interpreted cautiously and examined alongside the sensitivity analyses.

### 3.8 Small-study and reporting-bias assessment

The comparison-adjusted funnel plot included 31 comparison estimates (Fig. S2). Egger regression did not detect statistically significant funnel asymmetry (t = −0.96, 25 df; p = 0.346). This analysis was exploratory because asymmetry may arise from heterogeneity, multi-arm correlation or network geometry as well as selective reporting, and the non-significant result does not establish absence of reporting bias.

## Discussion

### 4.1 Principal findings

In this NMA of 28 RCTs contributing quantitative data, 14 adjunctive interventions had 95% CIs excluding the null versus the broad Control node. Mirtazapine, granisetron, tropisetron, minocycline and memantine occupied the five highest P-score positions, and tDCS, rTMS and body-oriented psychotherapy also showed favourable estimates. These findings extend earlier pairwise syntheses [7, 8] by placing pharmacological and non-pharmacological adjuncts in one connected framework.

The apparent hierarchy should not be read as a definitive ordering of clinical options. The primary network was predominantly a set of single-study active nodes connected to a pooled control, with few direct active–active comparisons. Consequently, large SMDs for the leading interventions may reflect genuine efficacy, sampling variability, scale properties or study-level differences. P-scores summarize relative point estimates and precision, but do not incorporate certainty, safety, acceptability or clinical importance [14].

### 4.2 Pharmacological add-on signals

Mirtazapine showed the largest primary estimate [41], while granisetron [49] and tropisetron [35] produced similarly large estimates. These agents act on serotonergic systems, and tropisetron additionally has cholinergic activity; however, the NMA estimates do not demonstrate that these mechanisms caused the observed clinical effects. Replication in adequately powered trials with harmonized negative-symptom phenotyping is needed before these signals can support routine prescribing.

Minocycline [32] and memantine [59] were also among the leading treatments, consistent with continued interest in inflammatory and glutamatergic pathways [3, 6]. The scale-specific PANSS-N estimate for minocycline was −4.65 points, but clinical importance cannot be inferred from this value alone because the network was sparse, follow-up was short and a generally accepted minimal important difference for this enriched population was not prespecified.

The very large effect size for mirtazapine (SMD −2.38) warrants particular caution. This magnitude is substantially larger than typical antipsychotic–placebo differences (SMD approximately 0.3–0.5) [9] and likely reflects the small sample size (n = 24 randomized) and potential instability of a single-study estimate rather than an exceptionally robust therapeutic effect.

### 4.3 Non-pharmacological add-on signals

tDCS [47] and rTMS trials [30, 39, 52, 61, 65] showed favourable estimates, as did body-oriented psychotherapy [24]. These interventions avoid additional medication exposure and may be attractive when polypharmacy or adverse effects are concerns. Nevertheless, stimulation parameters, treatment intensity and control conditions varied, and most comparisons were supported by small trials. The present analysis evaluated symptom efficacy rather than durability, functioning, acceptability or harms.

### 4.4 Network geometry, robustness and risk of bias

The low heterogeneity estimate (τ = 0.119) and stability of the five leading P-score positions after exclusion of flagged studies are reassuring but should be interpreted in light of only three heterogeneity degrees of freedom. The network had zero inconsistency degrees of freedom, so neither global decomposition nor node splitting was identifiable. Absence of an estimable inconsistency test is not evidence that consistency or transitivity holds.

Only eight trials were judged at low overall risk of bias; 15 had some concerns and five were high risk. Incomplete allocation-concealment reporting, attrition and unavailable prospective analysis specifications were recurring issues. The comparison-adjusted funnel plot and Egger test did not identify statistically detectable asymmetry, but power was limited and network geometry can obscure or mimic small-study effects. The large effects from several single-study nodes therefore require independent confirmation.

### 4.5 Strengths and limitations

Strengths include restriction to randomized adjunctive designs in adults selected for clinically relevant negative symptoms, explicit verification of stable background antipsychotic treatment, scale-specific sensitivity analyses, preservation of multi-arm trial structure, formal RoB 2 assessment and transparent separation of disconnected networks.

Several limitations materially constrain interpretation. First, despite supplementation with Europe PMC, Semantic Scholar, Crossref and citation chasing, Embase and CENTRAL records could not be exported from verified authorized sessions. This limitation may have introduced selection bias and remains the greatest source of uncertainty in review completeness. Second, 49 RCTs met the clinical and design criteria, but only 28 contributed to the locked quantitative dataset; the 21 otherwise eligible studies without usable outcome data in the assembled extraction set represent a potential availability bias. Third, many studies were small and short (3– 52 weeks), and definitions of prominent, predominant or persistent negative symptoms varied. Secondary negative symptoms due to depression, positive symptoms, extrapyramidal effects or sedation were not uniformly excluded.

Fourth, the primary SMD network combined endpoint and change scores and pooled placebo, sham, supportive counselling and treatment as usual. Stratified analyses were directionally compatible but sparse, whereas granular comparator nodes fragmented the network. Fifth, the broad outcome scales do not fully capture contemporary five-domain constructs [1–4]. Sixth, no formal CINeMA or GRADE certainty-of-evidence assessment was undertaken. Finally, adverse events, discontinuation, functioning and quality of life were outside the current efficacy analysis, preventing benefit–risk comparisons.

## Conclusions

Mirtazapine, granisetron, tropisetron, minocycline and memantine, together with selected non-pharmacological interventions, showed favourable efficacy signals as add-on treatments for negative symptoms of schizophrenia. The sparse, star-shaped evidence network, limited direct comparisons, non-estimable inconsistency, frequent risk-of-bias concerns and incomplete quantitative coverage mean that these rankings are hypothesis-generating. Adequately powered head-to-head RCTs using contemporary negative-symptom constructs, stable background treatment, longer follow-up and functional and safety outcomes are required.

## Data Availability

Use of artificial intelligence-assisted technology

## Acknowledgements

The authors acknowledge the institutional support of the Sichuan Provincial Center for Mental Health and the Key Laboratory of Psychosomatic Medicine, Chinese Academy of Medical Sciences.

## Statements and Declarations

## Funding

The authors did not receive support from any organization for the submitted work.

## Competing interests

The authors have no relevant financial or non-financial interests to disclose.

## Author contributions

Conceptualization: Chenkai Yangyang, Jia Chen, Xiaoqiang Xiao, Yifan Li, Hongjun Du, Wenjiao Min and Xu Zhang; methodology: Chenkai Yangyang, Jia Chen, Xiaoqiang Xiao and Xu Zhang; literature search and screening: Chenkai Yangyang, Jia Chen and Xiaoqiang Xiao; data extraction and curation: Chenkai Yangyang, Jia Chen and Xiaoqiang Xiao; formal analysis and visualization: Chenkai Yangyang and Jia Chen; writing— original draft: Chenkai Yangyang, Jia Chen, Xiaoqiang Xiao, Yifan Li and Hongjun Du; writing—review and editing: all authors; supervision: Wenjiao Min and Xu Zhang. All authors read and approved the final manuscript.

## Ethics approval

Not applicable. This systematic review and meta-analysis used aggregate data from previously published studies and did not involve new recruitment or access to identifiable participant data.

## Consent to participate

Not applicable.

## Consent for publication

Not applicable.

## Data availability

All data supporting the findings of this study, including the structured extraction dataset, are available from the corresponding author on reasonable request.

## Code availability

The R code used for the network meta-analysis and production of the statistical figures is available from the corresponding author on reasonable request.

## Use of artificial intelligence-assisted technology

OpenAI Codex was used to assist with language editing, document assembly and consistency checking. All scientific content, analyses, citations and final decisions were reviewed and approved by the authors, who take responsibility for the integrity of the work.

---

**Fig. S1.**
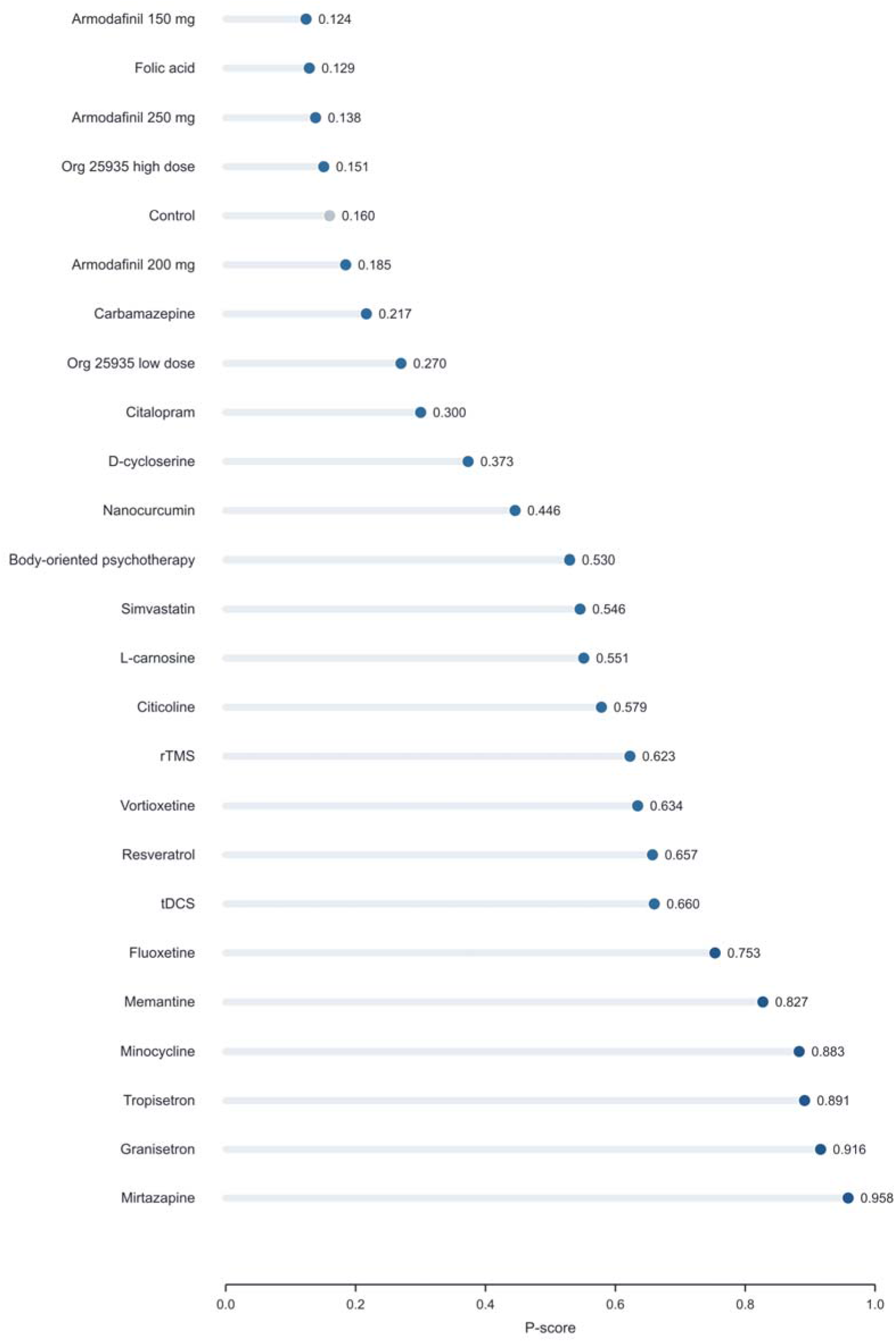
P-score ranking for the primary random-effects network. Higher values indicate a greater probability of being among the more effective treatments within this connected component; the ranking does not account for precision, certainty of evidence, safety or clinical importance

**Fig. S2.**
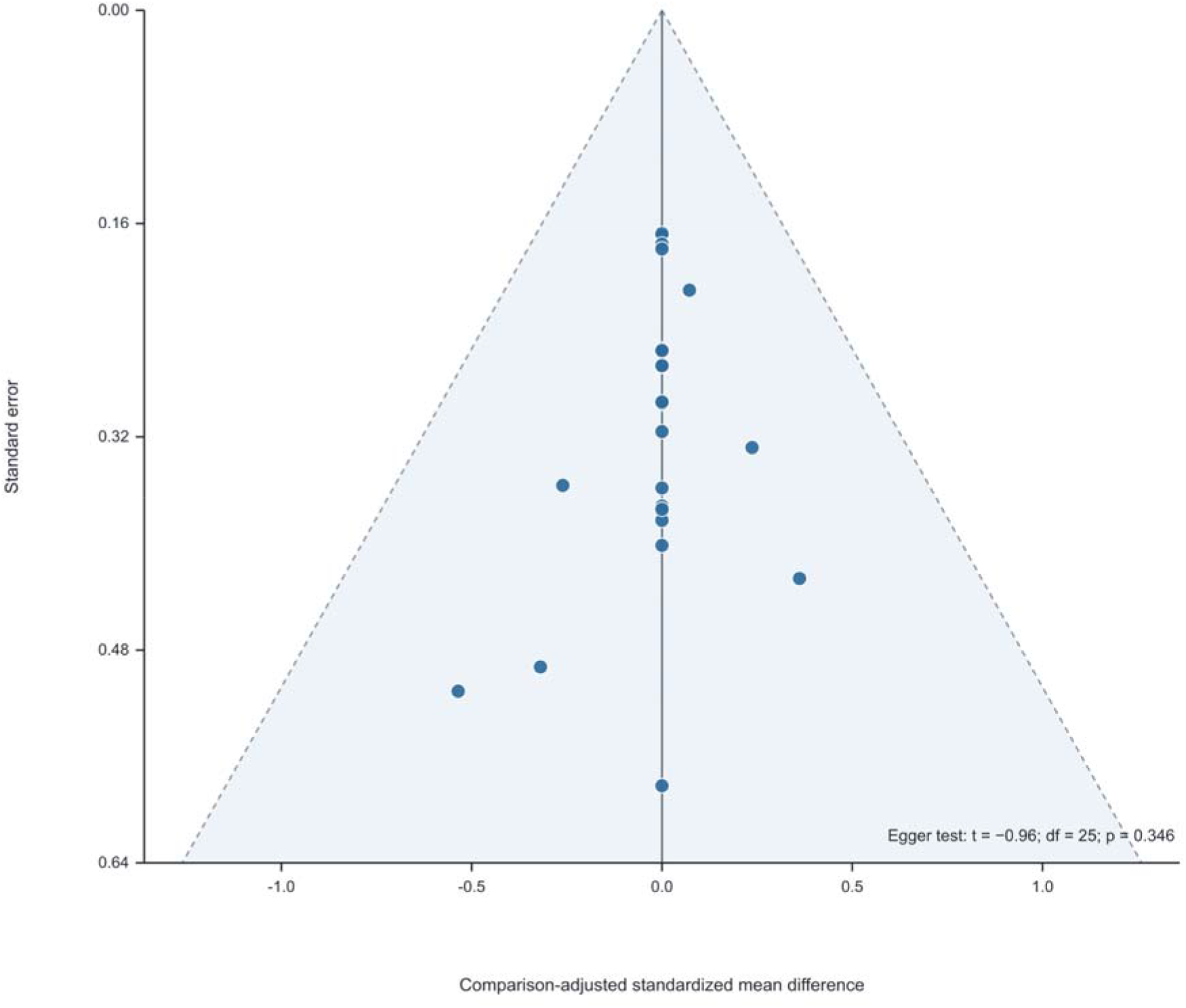
Comparison-adjusted funnel plot for the primary SMD network. The plot contains 31 comparison estimates. Egger regression gave t = −0.96 with 25 df (p = 0.346); the analysis is exploratory

## Notes

### Competing Interest Statement

The authors have declared no competing interest.

